# Post-Vaccination COVID-19 Infection and Its Correlates among Residents of Chandra, Gazipur, Bangladesh: A Cross-Sectional Study

**DOI:** 10.64898/2026.11.07.26367521

**Authors:** Abdullah Al Hasan Pranto, Syeda Sushmita Zafar, Mohammad Fakhruzzaman, Md. Safayeter Rahman, Sharmin Akter, Jannatul Taslima Meem, Rubaya Sultana, Shahid Afridi

## Abstract

**Background:** COVID-19 vaccines are highly effective at preventing severe disease and death but post-vaccination (breakthrough) infections are still seen, especially with immune-evasive variants. There is some evidence at the community level from Bangladesh. The aim of this study was to estimate prevalence, clinical profile and correlates of post-vaccination COVID-19 infection in Chandra, Kaliakair, Gazipur, Bangladesh.

**Methods:** This was a community-based cross-sectional study that included adults ≥18 years of age with at least one dose of a COVID-19 vaccine from March 2021 to December 2022. The respondents were 300 people who were selected by multistage sampling technique and interviewed with a pretested semi-structured questionnaire. Demographic, vaccination history, post-vaccination infection, clinical features, preventive behaviours, knowledge and comorbidities data were gathered. The chi-square test was used to test the bivariate associations and multivariable binary logistic regression was used to identify independent correlates. A p value of < 0.05 was used to determine statistical significance. Ethical Approval took from the IRB board of SCMST.

**Results:** Among 300 participants (mean age 48.0 ± 17.6 years; 52.0% male), 124 (41.3%) reported COVID-19 infection after vaccination. Of these, 24 (19.4%) were asymptomatic, 67 (54.0%) had mild disease, 24 (19.4%) had moderate disease, and 9 (7.3%) had severe disease; only 12 (9.7%) required hospitalisation. In multivariable analysis, rural residence (adjusted odds ratio [aOR] 4.87; 95% CI 2.42–9.79), poor mask use (aOR 2.28; 95% CI 1.31–3.97), poor physical distancing (aOR 2.74; 95% CI 1.55–4.86), poor hand hygiene (aOR 3.61; 95% CI 2.05–6.38), and diabetes (aOR 4.46; 95% CI 1.97–10.14) were independently associated with higher odds of infection, whereas receipt of a booster dose was protective (aOR 0.23; 95% CI 0.13–0.42).

**Conclusion:** Post-vaccination infection was common but largely mild and hospitalisation was rare. The booster vaccination was protective and factors such as rural residence, poor preventive practices and diabetes were risk factors for infection. These results do reinforce the importance of booster doses coupled with ongoing preventive behaviour and targeted interventions for clinically vulnerable populations.

## Introduction

The coronavirus disease 2019 (COVID-19) is a respiratory illness caused by a new type of coronavirus (CoV) called severe acute respiratory syndrome coronavirus 2 (SARS-CoV-2) that is highly transmissible. It was first reported in late 2019 and quickly became a global pandemic, and caused significant morbidity, mortality, and dysfunction in health, social and economic systems [1]. The clinical spectrum is from asymptomatic infection to fever, cough, sore throat, fatigue, loss of taste or smell, dyspnoea, pneumonia, respiratory failure and death [1,2].

Vaccination is one of the most powerful public health tools to help lower the burden of COVID-19 illness, hospitalisation and death. High level of protection has been shown in clinical trials and in real world studies, especially for severe disease and death [3–5] and large scale monitoring of vaccine performance is a priority. In February 2021, Bangladesh started a nationwide COVID-19 vaccination rollout, which began with frontline workers and older adults and people with underlying health conditions before opening to the general population [6]. However, vaccination immunity can fade over time and new variants can partially escape vaccine-induced immunity [5,7].

If a person tests positive for SARS-CoV-2 after being vaccinated against COVID-19, the infection is considered a post-vaccination COVID-19 infection, also known as vaccine breakthrough infection. A “breakthrough” case is defined as a laboratory confirmed case of infection that occurs at least 14 days after receiving the recommended primary vaccination series [8]. Breakthrough infection rates are also highly heterogeneous, depending on the type of vaccine, vaccine product, number of doses, time since vaccination, type of testing, occupational exposure, and the characteristics of the population [7–10]. Protection against hospitalisation and severe outcomes was still significant during the Omicron period, despite the increased frequency of breakthrough infections due to the higher transmissibility and immune-evasion properties of Omicron [5,7].

Vaccinated people who get infected have a reduced risk for developing severe illness and requiring a longer hospital stay compared to unvaccinated people [9,10]. However, fever, cough, headache, fatigue, sore throat, anosmia and dyspnoea can be present and hospitalization may be necessary, especially in older people and in individuals with comorbidities [9,10]. Various socio-demographic, clinical, behavioural and vaccination-related factors have been linked to post-vaccination COVID-19 infection, such as age, sex, occupation, education, family size, history of COVID-19 infection, comorbidity, vaccine type, incomplete vaccination, no booster dose, time since last dose, and contact with COVID-19 cases [7–10].

Immunisation is not a substitute for good preventive measures, particularly in the midst of active transmission. Wearing face mask, performing hand hygiene, ensuring adequate ventilation, avoiding close contact, practicing respiratory etiquette, testing after exposure, and isolating if ill all help reduce transmission [1,11]. In Bangladesh, studies have found variations in knowledge, attitudes, trust in vaccination and prevention behaviour based on education, age, place of living, profession, income and availability of trusted information [11–13].

While there is national and international data available, data at community level on post-vaccination COVID-19 infection and its correlates is limited, especially in Chandra, Gazipur. Local evidence is relevant since infection patterns and prevention practices can vary among different communities and work places. This study was thus designed to assess the COVID-19 infection after vaccination and its factors among the people of Chandra in Gazipur District. It included socio-demographic characteristics and vaccination history of the participants, estimates of prevalence and clinical characteristics of post vaccination infection, assessment of preventive health behaviours and vaccination related knowledge, and identification of factors statistically associated with infection. Results can be used to guide health education, booster dose messaging, risk reduction interventions, and future infectious disease preparedness efforts at the local level.

## Materials and Methods

The design of the study and setting in which it was conducted. The study was a cross-sectional study which was performed in Chandra community of Kaliakair Upazila, Gazipur District, Bangladesh from March 2021 to December 2022. The study period started in the wake of the national rollout of the COVID-19 vaccination campaign in February 2021, giving enough time for participants to be vaccinated at least once, and continued through the COVID-19 transmission waves of Delta and Omicron. A total of 20,395 individuals were selected for study, and their eligibility was determined. Adults (18 years old and older) who had received at least one dose of a COVID-19 vaccine and had been living in Chandra for at least six months prior to enrolment were included in the study. People who could not give informed consent or who were very sick during the time of the interview were excluded. Sample size and sampling The minimum required sample was approximately 92 subjects using standard cross-sectional formula n = Z2pq/d2, with the estimated prevalence of COVID-19 as 6.4%, 95% confidence level (Z = 1.96), and 5% margin of error (d). The sample was increased to 300 subjects to increase accuracy and allow subgroup analyses. This multistage sampling involved selection of unions randomly in the first stage and selection of households systematically in the second stage in the union of Chandra. An eligible adult from each household was invited to participate. Data collection instrument and procedure Based on an extensive literature search a semi-structured questionnaire was constructed, translated in Bangla and pretested with 30 participants who were not included in the final analysis. The socio-demographic characteristics, vaccination status (dose number, booster shots, type of vaccine and time gap from the last vaccination), clinical characteristics and severity of COVID-19 infection, preventive health behaviours, knowledge and attitudes toward COVID-19 vaccination, and comorbidities were collected using a questionnaire. Data was collected by using face-to-face interview using trained enumerators. Operational definitions A post-vaccination SARS-CoV-2 infection was defined as a laboratory confirmation of SARS-CoV-2 infection (either through PCR or rapid antigen test) at least 14 days after completing the recommended primary vaccination schedule, in line with the CDC definition [8]. Severity has been categorised as asymptomatic, mild (symptomatic but not breathless or requiring hospitalisation), moderate (requiring clinical assessment but not intensive care), or severe (requiring hospitalisation or a stay in intensive care). Participants were asked to self-report preventive behaviours (mask use, physical distancing, hand hygiene) as “sometimes/always” or “never/rarely”. Data handling and statistics Questionnaires were reviewed daily for completeness and consistency before coding, editing and entering into the Statistical Package for the Social Sciences (SPSS) version 26. Descriptive statistics were reported in terms of frequencies and percentages, means and standard deviations. Cross tabulations of the bivariate relationships between participant characteristics and post-vaccination infection were performed using the chi-square test. Variables with p < 0.10 in bivariate analysis and clinically relevant variables were put in a multivariable binary logistic regression model to look for independent correlates. Adjusted odds ratios (aORs) with 95% confidence intervals (CIs) were presented. Assessment of model calibration was done using the Hosmer– Lemeshow test, while the explanatory power was assessed using the Nagelkerke R^2^. Statistically significant results were considered those with p values of < 0.05. Ethical considerations All participants were given the objective and procedure of the study prior to the data collection and written informed consent was obtained. Participants were on a voluntary basis and confidentiality and anonymity were respected. Ethical clearance was obtained from the Institutional Ethical Review Committee (Approval No. SCMST/IRB/LAB/02-10/21-002, dated [10/02/2021]) before the start of the study and all procedures were followed as per the ethical guidelines of research involving human participants.

## Results

### Participant characteristics

A total of 300 vaccinated individuals were included in the study, with 156 (52.0%) males and 144 (48.0%) females. The mean age was 48.0 ± 17.6 years, and 92 (30.7%) participants were older than 60 years. Regarding residence, 94 (31.3%) lived in urban areas, 94 (31.3%) in semi-urban areas, and 112 (37.3%) in rural areas. Among respondents, 32.0% (96) had 1 dose, 32.0% (96) had 2 doses and 36.0% (108) had 3 doses of COVID-19 vaccine, and 46.3% (139) had received a booster dose. The average time gap between last vaccine dose and the survey was 18.5 ± 10.7 months (Table 1).

**Table 1.** Participant characteristics and post-vaccination outcomes (N = 300)

| Characteristic | Category | n (%) |
|---|---|---|
| Age | 18–30 years | 62 (20.7) |
|  | 31–45 years | 83 (27.7) |
|  | 46–60 years | 63 (21.0) |
|  | >60 years | 92 (30.7) |
| Gender | Male | 156 (52.0) |
|  | Female | 144 (48.0) |
| Residence | Urban | 94 (31.3) |
|  | Semi-urban | 94 (31.3) |
|  | Rural | 112 (37.3) |
| Vaccine doses | One | 96 (32.0) |
|  | Two | 96 (32.0) |
|  | Three | 108 (36.0) |
| Booster received | Yes | 139 (46.3) |
|  | No | 161 (53.7) |
| Post-vaccination infection | Yes | 124 (41.3) |
|  | No | 176 (58.7) |
| Infection severity<sup>1</sup> | Asymptomatic | 24 (19.4) |
|  | Mild | 67 (54.0) |
|  | Moderate | 24 (19.4) |
|  | Severe/other | 9 (7.3) |
| Hospitalisation<sup>1</sup> | Yes | 12 (9.7) |
|  | No | 112 (90.3) |
| Vaccination knowledge | Low | 16 (5.3) |
|  | Moderate | 153 (51.0) |
|  | High | 131 (43.7) |

<sup>1</sup>This was calculated based on 124 participants who had a post-vaccination infection.

### Prevalence and clinical characteristics of post-vaccination infection

124 participants (41.3%) reported post-vaccination COVID-19 infection while 176 (58.7%) did not report COVID-19 infection. Of these, 24 (19.4%) were asymptomatic, 67 (54.0%) had mild symptoms, 24 (19.4%) had moderate symptoms and 9 (7.3%) had severe or other recorded symptoms. Overall, 73.4% of the infected were either asymptomatic or suffering from mild disease. A total of 12 (9.7%) of the 124 infected participants needed hospitalisation while 112 (90.3%) were managed without hospitalisation (Table 1).

### Vaccination-related knowledge

In terms of COVID-19 vaccination knowledge, 16 (5.3%) participants scored low, 153 (51.0%) scored moderate and 131 (43.7%) scored high. The mean knowledge score was 15.8 ± 5.0 (Table 1).

### Factors associated with post-vaccination infection

The bivariate analysis revealed that residence, booster vaccination, the use of masks, hand hygiene, and diabetes were significant factors in post-vaccination COVID-19 infection (Table 2). The infection rate was 29.8% in urban areas, 36.2% in semi-urban areas, and 55.4% in rural areas (p < 0.001). Infection rates were higher among those without a booster dose versus those with a booster dose (51.6% vs 29.5%, p < 0.001). Non-use or infrequent use of masks was associated with increased risk (51.1% vs 33.1%; p = 0.002), as was poor hand hygiene (53.2% vs 30.8%; p < 0.001). Diabetics were more likely to be infected than non-diabetics (64.4% vs 37.3%; p = 0.001). Physical distancing had borderline association (p = 0.072) and hypertension had borderline association (p = 0.069).

**Table 2.** Bivariate associations with post-vaccination infection (N = 300)

| Factor | Category | Infected, n/N (%) | χ² | p-value |
|---|---|---|---|---|
| Residence | Urban | 28/94 (29.8) | 15.285 | <0.001 |
|  | Semi-urban | 34/94 (36.2) |  |  |
|  | Rural | 62/112 (55.4) |  |  |
| Booster received | No | 83/161 (51.6) | 14.070 | <0.001 |
|  | Yes | 41/139 (29.5) |  |  |
| Mask use | Never/rarely | 70/137 (51.1) | 9.181 | 0.002 |
|  | Sometimes/always | 54/163 (33.1) |  |  |
| Physical distancing | Never/rarely | 71/152 (46.7) | 3.238 | 0.072 |
|  | Sometimes/always | 53/148 (35.8) |  |  |
| Hand hygiene | Never/rarely | 75/141 (53.2) | 14.518 | <0.001 |
|  | Sometimes/always | 49/159 (30.8) |  |  |
| Diabetes | No | 95/255 (37.3) | 10.567 | 0.001 |
|  | Yes | 29/45 (64.4) |  |  |
| Hypertension | No | 88/230 (38.3) | 3.314 | 0.069 |
|  | Yes | 36/70 (51.4) |  |  |

Pearson’s chi-square test was used, and p < 0.05 was deemed significant. Only variables with a significant or close to significant p value are shown.

### Multivariable analysis

Multivariable logistic regression (Table 3) showed that living in a rural area was independently associated with an increased risk of infection over living in an urban area (aOR 4.87; 95% CI 2.42–9.79; p < 0.001). Poor mask use (aOR 2.28; 95% CI 1.31–3.97; p = 0.003), poor physical distancing (aOR 2.74; 95% CI 1.55–4.86; p = 0.001), and poor hand hygiene (aOR 3.61; 95% CI 2.05–6.38; p < 0.001) were independently associated with increased odds of infection. Diabetes was also found to be an independent risk factor (aOR 4.46; 95% CI 1.97–10.14; p < 0.001), while there was a protective effect from receiving a booster dose (aOR 0.23; 95% CI 0.13–0.42; p < 0.001), which equates to around 77% lower odds of infection. After adjustment, age >60 years, semi-urban residence, and hypertension were not significantly associated with infection. The model was well calibrated (Hosmer– Lemeshow p = 0.707) and accounted for about 35.5% of the variance in the outcome (Nagelkerke R^2^ = 0.355).

**Table 3.** Multivariable logistic regression of factors associated with post-vaccination infection.

| Predictor | aOR | 95% CI | p-value |
|---|---|---|---|
| Age >60 years | 1.51 | 0.81–2.83 | 0.194 |
| Semi-urban residence | 1.45 | 0.71–2.96 | 0.302 |
| Rural residence | 4.87 | 2.42–9.79 | <0.001 |
| Poor mask use | 2.28 | 1.31–3.97 | 0.003 |
| Poor physical distancing | 2.74 | 1.55–4.86 | 0.001 |
| Poor hand hygiene | 3.61 | 2.05–6.38 | <0.001 |
| Diabetes | 4.46 | 1.97–10.14 | <0.001 |
| Hypertension | 1.35 | 0.69–2.65 | 0.378 |
| Booster received | 0.23 | 0.13–0.42 | <0.001 |

Adjusted odds ratio (aOR); CI: confidence interval. The reference categories are: age < or = 60 years, urban residents, appropriate preventive behaviour, lack of the corresponding morbidity, and no booster dose. Model fit: Hosmer–Lemeshow p = 0.707; Nagelkerke R^2^ = 0.355.

## Discussion

This study reports that 41.3% of the subjects were infected with COVID-19 even after vaccination. While this percentage may seem high, the rates of breakthrough infection are highly variable across different populations, variants, periods of follow-up time, vaccination types, testing capacity, and time since vaccination [5,7]. SARS-CoV-2 immunity is transient and is weaker against variants like Omicron that are less susceptible to immune responses, while immunity against severe disease and hospitalisation remains relatively strong [5,7]. In line with this, 73.4% of infected respondents in the current study experienced either no symptoms or mild symptoms and 9.7% were hospitalised. This finding is similar to the report from Krishna et al., where the majority of the breakthrough infections in the vaccinated healthcare workers were mild, which led to the conclusion of the benefit of vaccination in preventing severe clinical outcomes [14]. However, it is necessary to be careful in comparing prevalence estimates across studies in that some studies may have only included people who were fully vaccinated with the primary series while the current sample had a mix of doses. Variations between studies are due to differences in diagnostic confirmation, recall periods, and exposure. All these observations indicate that although vaccination decreases the severity of SARS-CoV-2 infection, it is not fully protective against infection.

The current analysis showed that rural residence, non-compliance for mask use, poor physical distancing, inadequate hand hygiene, and diabetes were associated with an increased chance of getting an infection after COVID-19 vaccination. The risk of infection for the rural group was about 4.87 times higher than for the urban group. Such association might be due to differences in available reliable health information, access to diagnostic services, booster vaccination, occupation exposure and adherence to preventive recommendations. Poor mask use, poor physical distancing and poor hand hygiene were all individually associated with a higher risk, suggesting that vaccination, combined with consistent adherence to masking, physical distancing and hand hygiene is most effective, especially during periods of high community transmission. Previously reported similar findings and an inverse relationship between severity of post vaccination disease and compliance with preventive measures has been reported [15]. Diabetics had around a 4.46-fold increased chance of infection. Metabolic and immune impairment associated with diabetes can contribute to susceptibility, reduce immune functions and pre-dispose to negative COVID-19 outcomes. However, receiving a booster dose was protective, with the odds of infection decreasing by ∼77% following adjustment. This is in line with findings that immunity wanes after booster dose series, especially against new variants [5,7]. Older age (over 60 years) and hypertension were no longer significant risk factors for infection after adjustment, possibly reflecting the indirect effects of other clinical and behavioral factors. The model was well calibrated, and accounted for 35.5% of the variance in the outcomes, indicating that other risk factors, such as previous infection, viral variant, occupation, household crowding, and immune status, may also be involved. This study has a number of limitations. First, because it is a cross sectional study, temporality of exposures and infection cannot be established. Second, the infection status and preventive behaviours were self-reported and may be prone to recall and social desirability biases. Third, no data on circulating variants, previous infection or comprehensive immune profiles were available. Fourth, the type of vaccine received was documented, but was not analysed separately due to low subgroup sizes. Fifth, the study took place in one upazila, and so these findings might not be generalized to other upazilas in Bangladesh. Last, there may be confounding of other factors that could not be measured, including household crowding and occupational exposure. Laboratory confirmation of future prospective studies with characterization of the variants is indicated.

## Conclusion

Among the 41.3% of those vaccinated who reported a COVID-19 infection after vaccination, the vast majority of infections were asymptomatic or mild and hospitalization was rare. Being rural, not using a mask well, not maintaining good physical distancing, not cleaning their hands well, and having diabetes were risk factors for infection, while booster vaccination was significantly protective. These findings underscore the need for synergies between vaccination and booster doses, and for the promotion of ongoing prevention practices, as well as the need for targeted support for clinically vulnerable groups. Time of infection, variant, occupational exposure, previous infection and immune status should be studied in future studies using prospective community data.

## Data Availability

All data produced in the present work are contained in the manuscript

## Data availability

The dataset analyzed during the present study is available from the corresponding author on reasonable request.

## Ethical approval

Ethical clearance was obtained from the Institutional Ethical Review Committee of Saic College of Medical Science and Technology (SCMST) (Approval No. SCMST/IRB/LAB/02-10/21-002, dated [10/02/2021]) prior to the commencement of the study.

## Author Contributions

**Abdullah Al Hasan Pranto** — GROUP 1: Conception of the work, Acquisition and Analysis of data; GROUP 2: Revising the work critically for important intellectual content; GROUP 3: Final approval of the version to be published; GROUP 4: Agree to be accountable for all aspects of the work in ensuring that questions related to the accuracy or integrity of any part of the work are appropriately investigated and resolved.

**Dr. Syeda Sushmita Zafar** — GROUP 1: Design of the work, Analysis and Interpretation of data; GROUP 2: Revising the work critically for important intellectual content; GROUP 3: Final approval of the version to be published; GROUP 4: Agree to be accountable for all aspects of the work.

**Dr. Mohammad Fakhruzzaman** — GROUP 1: Design of the work, Analysis and Interpretation of data; GROUP 2: Revising the work critically for important intellectual content; GROUP 3: Final approval of the version to be published; GROUP 4: Agree to be accountable for all aspects of the work.

**Md. Safayeter Rahman** — GROUP 1: Analysis and Interpretation of data; GROUP 2: Revising the work critically for important intellectual content; GROUP 3: Agree to be accountable for all aspects of the work.

**Sharmin Akter** — GROUP 1: Interpretation of data; GROUP 2: Revising the work critically for important intellectual content; GROUP 3: Final approval of the version to be published.

**Jannatul Taslima Meem** — GROUP 1: Analysis of data; GROUP 2: Revising the work critically for important intellectual content; GROUP 3: Final approval of the version to be published.

**Rubaya Sultana** — GROUP 1: Acquisition of data, Analysis and Interpretation of data; GROUP 2: Revising the work critically for important intellectual content; GROUP 3: Final approval of the version to be published.

**Shahid Afridi** — GROUP 1: Analysis and Interpretation of data; GROUP 2: Revising the work critically for important intellectual content; GROUP 3: Final approval of the version to be published.

